# Evaluation and Improvement of the National Early Warning Score (NEWS2) for COVID-19: a multi-hospital study

**DOI:** 10.1101/2020.04.24.20078006

**Authors:** Ewan Carr, Rebecca Bendayan, Daniel Bean, Matt Stammers, Wenjuan Wang, Huayu Zhang, Thomas Searle, Zeljko Kraljevic, Anthony Shek, Hang T T Phan, Walter Muruet, Rishi K Gupta, Anthony J Shinton, Mike Wyatt, Ting Shi, Xin Zhang, Andrew Pickles, Daniel Stahl, Rosita Zakeri, Mahdad Noursadeghi, Kevin O’Gallagher, Matt Rogers, Amos Folarin, Christopher Bourdeaux, Chris McWilliams, Lukasz Roguski, Florina Borca, James Batchelor, Xiaodong Wu, Jiaxing Sun, Ashwin Pinto, Bruce Guthrie, Cormac Breen, Abdel Douiri, Honghan Wu, Vasa Curcin, James T Teo, Ajay M Shah, Richard J B Dobson

**Author notes:** **Corresponding author:** Dr Ewan Carr, Institute of Psychiatry, Psychology & Neuroscience (IoPPN), 16 De Crespigny Park, London, SE5 8AF. Joint First Author (Ewan Carr, Rebecca Bendayan). Joint Last Author (James T Teo, Ajay M Shah, Richard J B Dobson).

## Abstract

**Background:** The National Early Warning Score (NEWS2) is currently recommended in the United Kingdom for risk stratification of COVID outcomes, but little is known about its ability to detect severe cases. We aimed to evaluate NEWS2 for severe COVID outcome and identify and validate a set of routinely-collected blood and physiological parameters taken at hospital admission to improve the score.

**Methods:** Training cohorts comprised 1276 patients admitted to King’s College Hospital NHS Foundation Trust with COVID-19 disease from 1^st^ March to 30^th^ April 2020. External validation cohorts included 5037 patients from four UK NHS Trusts (Guys and St Thomas’ Hospitals, University Hospitals Southampton, University Hospitals Bristol and Weston NHS Foundation Trust, University College London Hospitals), and two hospitals in Wuhan, China (Wuhan Sixth Hospital and Taikang Tongji Hospital). The outcome was severe COVID disease (transfer to intensive care unit or death) at 14 days after hospital admission. Age, physiological measures, blood biomarkers, sex, ethnicity and comorbidities (hypertension, diabetes, cardiovascular, respiratory and kidney diseases) measured at hospital admission were considered in the models.

**Results:** A baseline model of ‘NEWS2 + age’ had poor-to-moderate discrimination for severe COVID infection at 14 days (AUC in training sample = 0.700; 95% CI: 0.680, 0.722; Brier score = 0.192; 95% CI: 0.186, 0.197). A supplemented model adding eight routinely-collected blood and physiological parameters (supplemental oxygen flow rate, urea, age, oxygen saturation, CRP, estimated GFR, neutrophil count, neutrophil/lymphocyte ratio) improved discrimination (AUC = 0.735; 95% CI: 0.715, 0.757) and these improvements were replicated across five UK and non-UK sites. However, there was evidence of miscalibration with the model tending to underestimate risks in most sites.

**Conclusions:** NEWS2 score had poor-to-moderate discrimination for medium-term COVID outcome which raises questions about its use as a screening tool at hospital admission. Risk stratification was improved by including readily available blood and physiological parameters measured at hospital admission, but there was evidence of miscalibration in external sites. This highlights the need for a better understanding of the use of early warning scores for COVID.

**Key messages:**

- The National Early Warning Score (NEWS2), currently recommended for stratification of severe COVID-19 disease in the UK, showed poor-to-moderate discrimination for medium-term outcomes (14-day transfer to ICU or death) among COVID-19 patients.
- Risk stratification was improved by the addition of routinely-measured blood and physiological parameters routinely at hospital admission (supplemental oxygen, urea, oxygen saturation, CRP, estimated GFR, neutrophil count, neutrophil/lymphocyte ratio) which provided moderate improvements in a risk stratification model for 14-day ICU/death.
- This improvement over NEWS2 alone was maintained across multiple hospital trusts but the model tended to be miscalibrated with risks of severe outcomes underestimated in most sites.
- We benefited from existing pipelines for informatics at KCH such as CogStack that allowed rapid extraction and processing of electronic health records. This methodological approach provided rapid insights and allowed us to overcome the complications associated with slow data centralisation approaches.

## Background

As of 29^th^ September 2020, there have been >33 million confirmed cases of COVID-19 disease worldwide(1). While approximately 80% of infected individuals have mild or no symptoms(2), some develop severe COVID-19 disease requiring hospital admission. Within the subset of those requiring hospitalisation, early identification of those who deteriorate and require transfer to an intensive care unit (ICU) for organ support or may die is vital.

Currently available risk scores for deterioration of acutely-ill patients include (i) widely-used generic ward-based risk indices such as the National Early Warning Score (NEWS2)(3), (ii) the Modified Sequential Organ Failure Assessment (mSOFA)(4) and Quick Sequential Organ Failure Assessment(5) scoring systems; and (iii) the pneumonia-specific risk index, CURB-65(6) which usefully combines physiological observations with limited blood markers and comorbidities.

NEWS2 is a summary score of six physiological parameters or ‘vital signs’ (respiratory rate, oxygen saturation, systolic blood pressure, heart rate, level of consciousness, temperature and supplemental oxygen dependency), used to identify patients at risk of early clinical deterioration in the United Kingdom (UK) NHS hospitals(7,8) and primary care. The physiological parameters assessed in the NEWS2 score (in particular, patient temperature, oxygen saturation and supplemental oxygen dependency) have previously been associated with COVID-19 outcomes(2), but little is known about their predictive value for COVID-19 disease severity in hospitalised patients(9). Additionally, a number of COVID-19-specific risk indices are being developed(10,11) as well as unvalidated online calculators(12) but generalisability is unknown(13). A Chinese study has suggested a modified version of NEWS2 with addition of age only(14) but without any data on performance. With near universal usage of NEWS2 in UK NHS Trusts since March 2019(15), a minor adaptation to NEWS2 would be relatively easy to implement.

As the SARS-Cov2 pandemic has progressed there has been a growing body of research aiming to develop risk prediction models to support clinical decisions, triage and care in hospitalised patients(13). Within this context, evidence has emerged regarding potentially useful blood biomarkers(2,16–19). Although most early reports contained data from small numbers of patients, several markers have consistently been associated with severe outcomes. These include neutrophilia and lymphopenia, particularly in older adults(11,18,20,21), neutrophil-to-lymphocyte ratio(22), C-reactive Protein (CRP) and lymphocyte-to-CRP ratio(22), markers of liver and cardiac injury such as alanine aminotransferase (ALT), aspartate aminotransferase (AST) and cardiac troponin(23) and elevated D-dimers, ferritin and fibrinogen (2,6,8).

A recent systematic review identified CRP and creatinine as common predictors of COVID severity and mortality(13). However, this review found most existing studies to be at high risk of bias due to non-representative samples, model overfitting, or poor reporting. Many sampling issues stem from the rapid development of prediction models using initial cohorts of COVID patients, often from a single site without external validation. Furthermore there was a lack of community testing which prevented comparisons to traditional control groups. In the present study we present a cross-cohort analysis that builds upon our preliminary work(24) which suggested that adding age and common blood biomarkers to the NEWS2 score could improve risk prediction in patients hospitalized with COVID. While incorporating external validation, this preliminary work was limited in that the training sample comprised 439 patients (the cohort available at the time of model development). In the present study we build on this preliminary work by (i) expanding the cohort used for model development to all 1276 patients at KCH; (ii) using hospital admission (rather than symptom onset) as the index date; (iii) considering in sensitivity analyses shorter-term (3-day) outcomes; (iv) improving the reporting of model calibration and clinical utility in validation sites; and (v) increasing the number of external sites.

Thus, our aim is to evaluate the NEWS2 score and identify which clinical and blood biomarkers routinely measured at hospital admission can improve medium-term risk stratification of severe COVID outcome at 14 days from hospital admission. Our specific objectives were:

1. To explore independent associations of routinely measured physiological and blood parameters (including NEWS2 parameters) at hospital admission with disease severity (ICU admission or death at 14 days from hospital admission), adjusting for demographics and comorbidities;
2. To develop a prediction model for severe COVID outcomes at 14 days combining multiple blood and physiological parameters.
3. To compare the predictive value of the resulting model with NEWS2 score alone using (i) internal validation; (ii) external validation at five hospital sites.

## Methods

### Study cohorts

The KCH training cohort (n=1276) was defined as all adult inpatients testing positive for SARS-Cov2 by reverse transcription polymerase chain reaction (RT-PCR) between 1^st^ March to 31^st^ April 2020 at two acute hospitals (King’s College Hospital and Princess Royal University Hospital) in South East London (UK) of Kings College Hospital NHS Foundation Trust (KCH). All patients included in the study had symptoms consistent with COVID-19 (e.g. cough, fever, dyspnoea, myalgia, delirium, diarrhoea). For external validation purposes we used five cohorts:

1. Guys and St Thomas’ Hospital NHS Foundation Trust (GSTT) of 988 cases (3^rd^ March 2020 to 26^th^ August 2020)
2. University Hospitals Southampton NHS Foundation Trust (UHS) of 633 cases (7^th^ March to 6th June 2020)
3. University Hospitals Bristol and Weston NHS Foundation Trust (UHBW) of 190 cases (12^th^ March to 11^th^ June 2020)
4. University College Hospital London (UCH) of 411 cases (1st February to 30th April 2020).
5. Wuhan Sixth Hospital and Taikang Tongji Hospital of 2815 cases (4^th^ February 2020 to 30th March 2020)

Data were extracted from structured and/or unstructured components of electronic health records (EHR) in each site. Details regarding data processing and ethics at each site are presented in Supplementary Materials.

### Measures

#### Outcome

For all sites, the outcome was severe COVID disease at 14 days following hospital admission, categorised as transfer to ICU/death (WHO-COVID-19 Outcomes Scales 6-8) vs. not transferred to ICU/death (Scales 3-5). For nosocomial patients (patients with symptom onset after hospital admission) the endpoint was defined as 14 days after symptom onset. Dates of hospital admission, symptom onset, ICU transfer and death were extracted from electronic health records or ascertained manually by a clinician.

#### Blood and physiological parameters

We included blood and physiological parameters that were routinely obtained at hospital admission which are routinely available in a wide range of national and international hospital and community settings. Measures available for fewer than 30% of patients were not considered (including Troponin-T, Ferritin, D-dimers and HbA1c, GCS score). We excluded creatinine since this parameter correlates highly (*r* > 0.8) with, and is used in the derivation of, estimated GFR. We excluded white blood cell count (WBCs) which is highly correlated with neutrophil and lymphocyte counts.

The candidate blood parameters therefore comprised: albumin (g/L), C-reactive protein (CRP; mg/L), estimated Glomerular Filtration Rate (eGFR; mL/min), Haemoglobin (g/L), lymphocyte count (x 10^9^/L), neutrophil count (x 10^9^/L), and platelet count (PLT; x 10^9^/L), neutrophil-to-lymphocyte ratio (NLR), lymphocyte-to-CRP ratio[21], and urea (units). The candidate physiological parameters included the NEWS2 total score, as well as the following parameters: respiratory rate (breaths per minute), oxygen saturation (%), supplemental oxygen flow rate (L/min), diastolic blood pressure (units), systolic blood pressure (mmHg), heart rate (beats/min), temperature (°C), and consciousness (Glasgow Coma Scale; GCS). For all parameters we used the first available measure up to 48 hours following hospital admission.

#### Demographics and comorbidities

Age, sex, ethnicity and comorbidities were considered. Self-defined ethnicity was categorised as White vs. non-White (Black, Asian, and minority ethnic) and patients with ethnicity recorded as ‘unknown/mixed/other’ were excluded (n=316; 25%). Binary variables were derived for comorbidities: hypertension, diabetes, heart disease (heart failure and ischemic heart disease), respiratory disease (asthma and chronic obstructive pulmonary disease, COPD) and chronic kidney disease.

### Statistical analyses

All continuous parameters were winsorized (at 1% and 99%) and scaled (mean = 0; standard deviation = 1) to facilitate interpretability and comparability(25). Logarithmic or square-root transformations were applied to skewed parameters. To explore independent associations of blood and physiological parameters with 14-day ICU/death (Objective 1) we used logistic regression with Firth’s bias reduction method(26). Each parameter was tested independently, adjusted for age and sex (Model 1) and then additionally adjusted for comorbidities (Model 2). *P*-values were adjusted using the Benjamini-Hochberg procedure to keep the False Discovery Rate (FDR) at 5%(27).

To evaluate NEWS2 and identify parameters that could improve prediction of severe COVID outcomes (Objectives 2 and 3) we used regularized logistic regression with a LASSO (Least Absolute Shrinkage and Selection Operator) estimator that shrinks parameters according to their variance, reduces overfitting, and enables automatic variable selection(28). The optimal degree of regularization was determined by identifying a tuning parameter λ using cross-validation. To avoid overfitting and to reduce the number of false positive predictors, λ was selected to give a model with area under the curve (AUC) one standard error below the ‘best’ model. To evaluate the predictive performance of our model on new cases of the same underlying population (internal validation), we performed nested cross-validation (10 folds for inner loop; 10 folds/1000 repeats for outer loop). Discrimination was assessed using AUC and Brier score. Missing feature information was imputed using k-Nearest Neighbours imputation (k=5). All steps (feature selection, winsorizing, scaling, and kNN imputation) were incorporated within the model development and selection process to avoid data leakage that would otherwise result in optimistic performance measures(29). All analyses were conducted with Python 3.8 (30) using the statsmodels(31) and Scikit-Learn(32) packages.

We evaluated the transportability of the derived regularized logistic regression model in external validation samples from GSTT (n=988), UHS (n=564), UHBW (n=190), UCH (n=411), and Wuhan (n=2815). Validation used LASSO logistic regression models trained on the KCH training sample, with code and pre-trained models shared via GitHub^1^. Models were assessed in terms of discrimination (AUC, sensitivity, specificity, Brier score), calibration, and clinical utility (decision curve analysis)(25). Moderate calibration was assessed by plotting model predicted probabilities (x-axis) against observed proportions (y-axis) with LOESS logistic curves(33). Clinical utility was assessed using decision curve analysis where ‘net benefit’ was plotted against a range of threshold probabilities. Unlike diagnostic performance measures, decision curves incorporate preferences of the clinician and patient. The threshold probability (p_t_) is where the expected benefit of treatment is equal to the expected benefit of avoiding treatment(34). Net benefit was calculated by counting the number of true positives (predicted risk > p_t_ and experienced severe COVID outcome) and false positives (predicted risk > p_t_ but did not experience severe COVID outcome), and using the below formula:

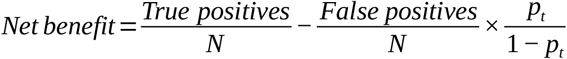

Our model was developed as a screening tool, to identify at hospital admission those patients who were at risk of more severe outcomes. The intended treatment for patients with a positive result from this model would be further examination by a clinician, who would make recommendations regarding appropriate treatment (e.g. earlier transfer to ICU, intensive monitoring, treatment). We compared the decision curve from our model to two extreme cases of ‘treat none’ and ‘treat all’. The ‘treat none’ (i.e. routine management) strategy implies that no patients would be selected for further examination by a clinician; the ‘treat all’ strategy (i.e. intensive management) implies that all patients would undergo further assessment. A model is clinically beneficial if the model-implied net benefit is greater than either the ‘treat none’ or ‘treat all’ strategies.

Since the intended strategy involves further examination by a clinician, and is therefore low risk, our emphasis throughout is on avoiding false negatives (i.e. failing to detect a severe case) at the expense of false positives. We therefore used thresholds of 30% and 20% (for 14-day and 3-day outcomes, respectively) to calculate sensitivity and specificity. This gave a better balance of sensitivity vs. specificity and reflected the clinical preference to avoid false negatives for the proposed screening tool.

#### Sensitivity analyses

We conducted four sensitivity analyses. First, to explore the ability of NEWS2 to predict shorter-term severe COVID outcome we developed models for ICU transfer/death at 3 days following hospital admission. All steps described above were repeated, including training (feature selection) and external validation. Second, following recent studies suggesting sex differences in COVID outcome(18) we tested interactions between each physiological and blood parameter and sex using likelihood-ratio tests. Third, we repeated all models with adjustment for ethnicity in the subset of individuals with available data for ethnicity (n=960 in the KCH training sample). Finally, to explore differences between community-acquired vs. nosocomial infection, we repeated all models after excluding 153 nosocomial patients (n=1123).

## Results

### Descriptive analyses

The KCH training cohort comprised 1276 patients admitted with a confirmed diagnosis of COVID-19 (from 1^st^ March to 31^st^ March 2020) of whom 389 (31%) and were transferred to ICU or died within 14 days of hospital admission, respectively. The validation cohorts comprised 5037 patients across five sites. At UK NHS trusts, 30% to 42% of patients were transferred to ICU or died within 14 days of admission. Disease severity was lower in the Wuhan sample, where 4% were transferred to ICU or died. Table 1 presents the demographic and clinical characteristics of the training and validation cohorts. The UK sites were similar in terms of age and sex, with patients tending to be older (median age 66-74) and male (58% to 63%), but varied in the proprotion of patients of non-White ethnicity (from 10% at UHS to 40% at KCH and UCH). Blood and physiological parameters were broadly consistent across UK sites.

**Table 1:**
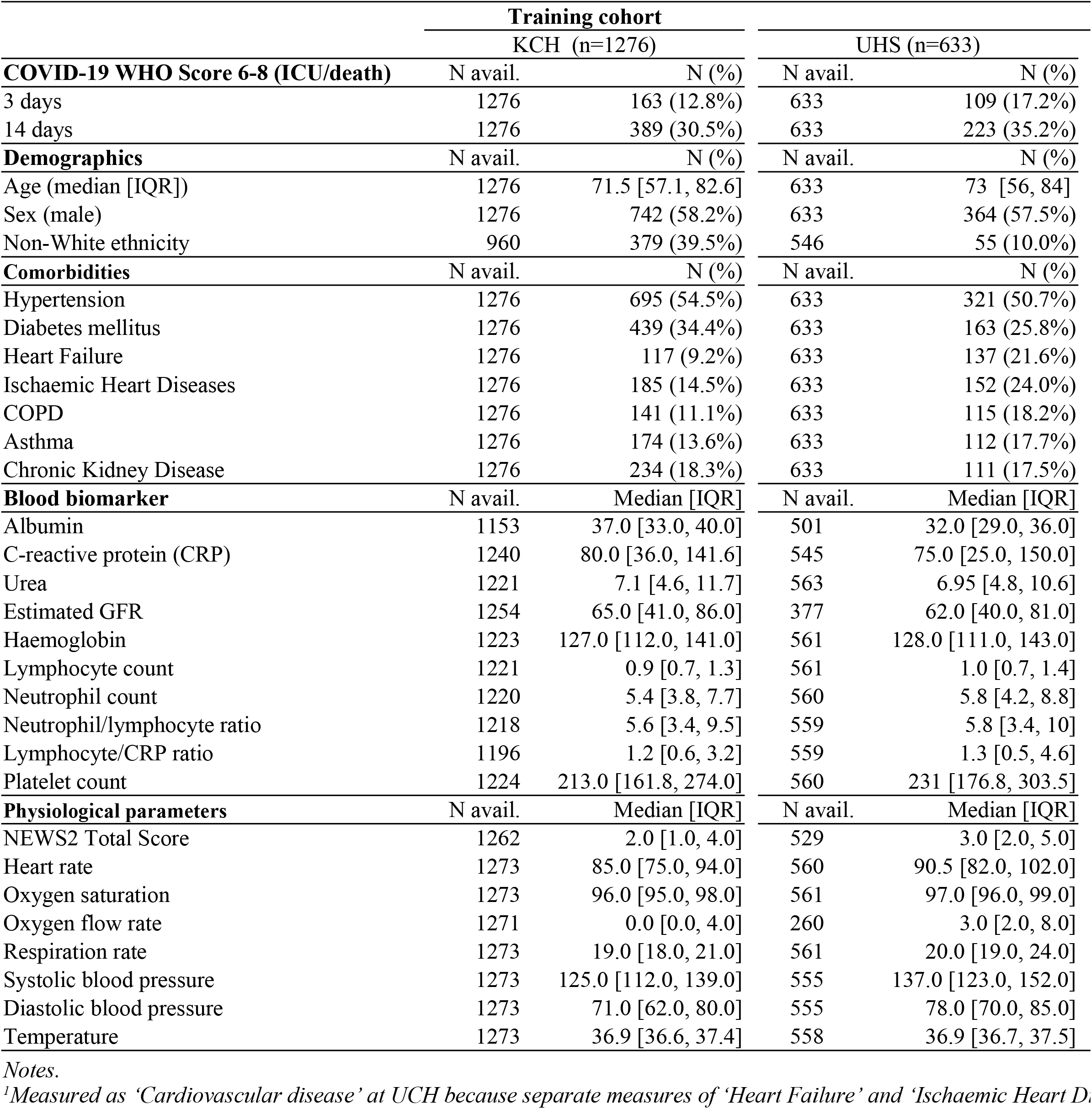
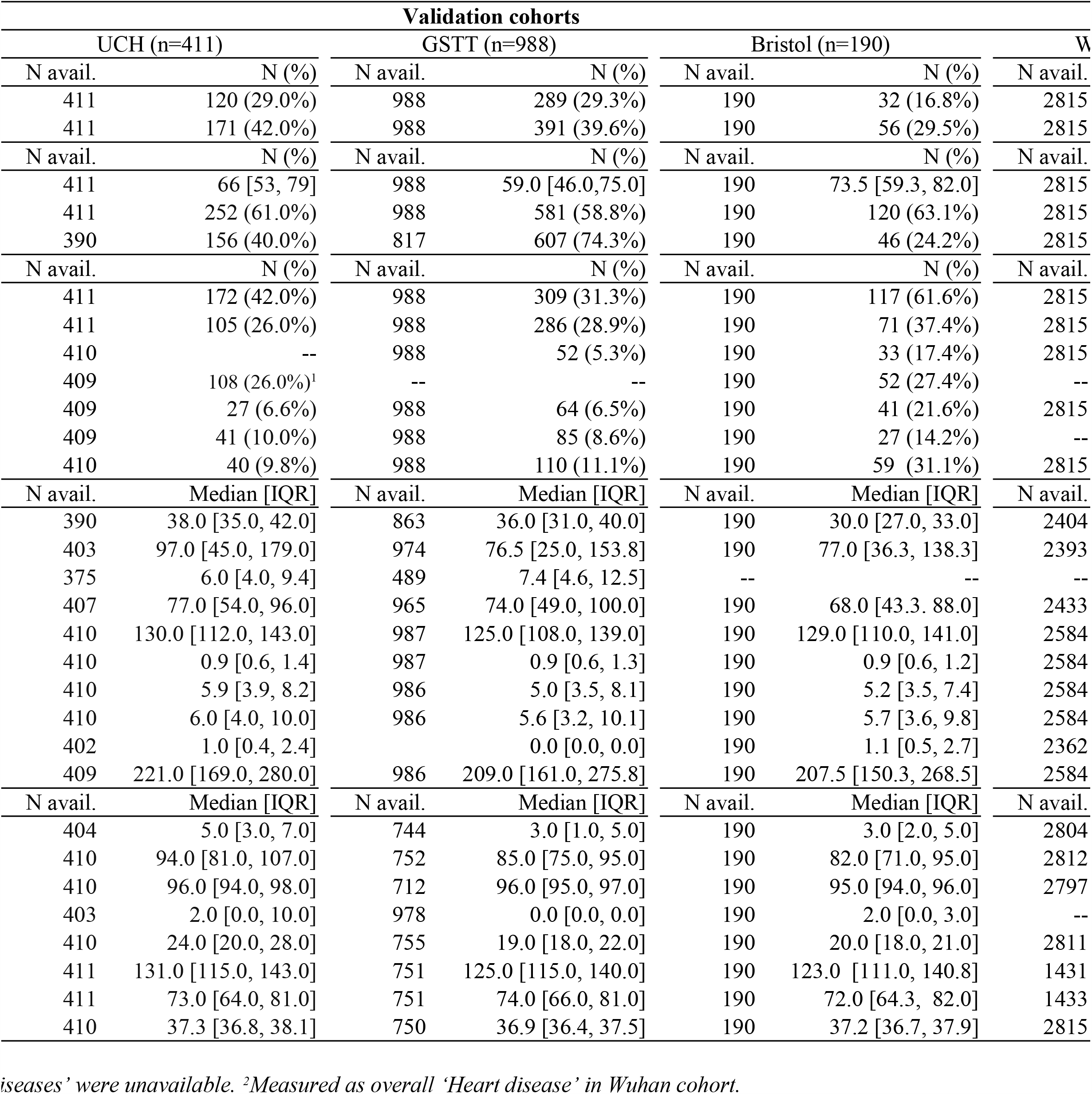
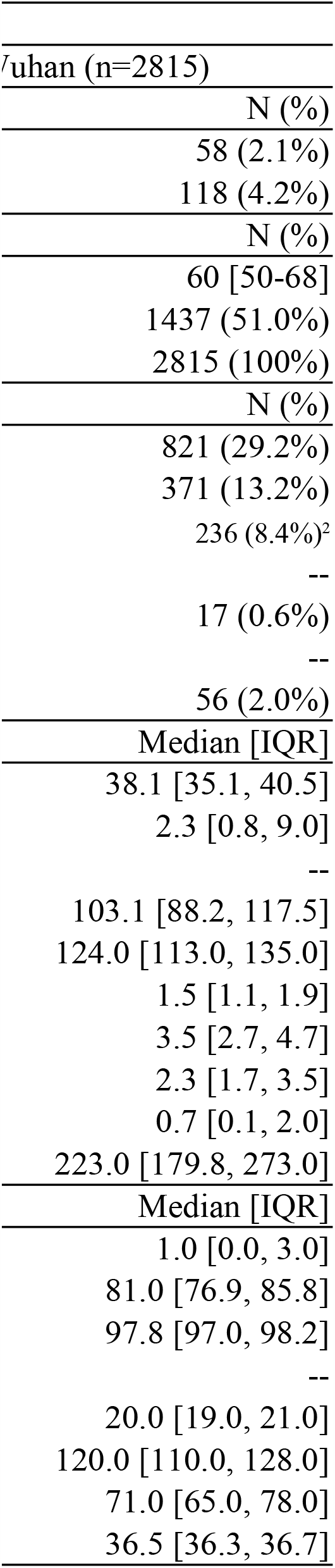
Patient characteristics of the training/validation cohorts

Logistic regression models were used to assess independent associations between each variable and severe COVID outcome (ICU transfer/death) in the KCH cohort. Supplementary Table 1 presents adjusted odds ratios adjusted for age and sex (Model 1) and comorbidities (Model 2), sorted by effect size. Increased odds of transfer to ICU or death by 14 days were associated with NEWS2 score, oxygen flow rate, respiratory rate, CRP, neutrophil count, urea, neutrophil/lymphocyte ratio, heart rate, and temperature. Reduced odds of severe outcomes were associated with lymphocyte/CRP ratio, oxygen saturation, estimated GFR, and Albumin.

#### Evaluating NEWS2 score for prediction of severe COVID outcome

Logistic regression models were used to evaluate a baseline model containing hospital admission NEWS2 score and age for prediction of severe COVID outcomes at 14 days. Internally validated discrimination for the KCH training sample was moderate (AUC = 0.700; 95% CI: 0.680, 0.722; Brier score = 0.192; 0.186, 0.197; Table 2). Discrimination remained poor-to-moderate in UK validation sites (AUC = 0.623 to 0.729) but good in Wuhan hospitals (AUC = 0.815; Figures 1 and 2). Calibration was inconsistent with risks underestimated in some sites (UHS, GSTT) and overestimated in others (UHBW) (Figure 2).

**Table 2:**
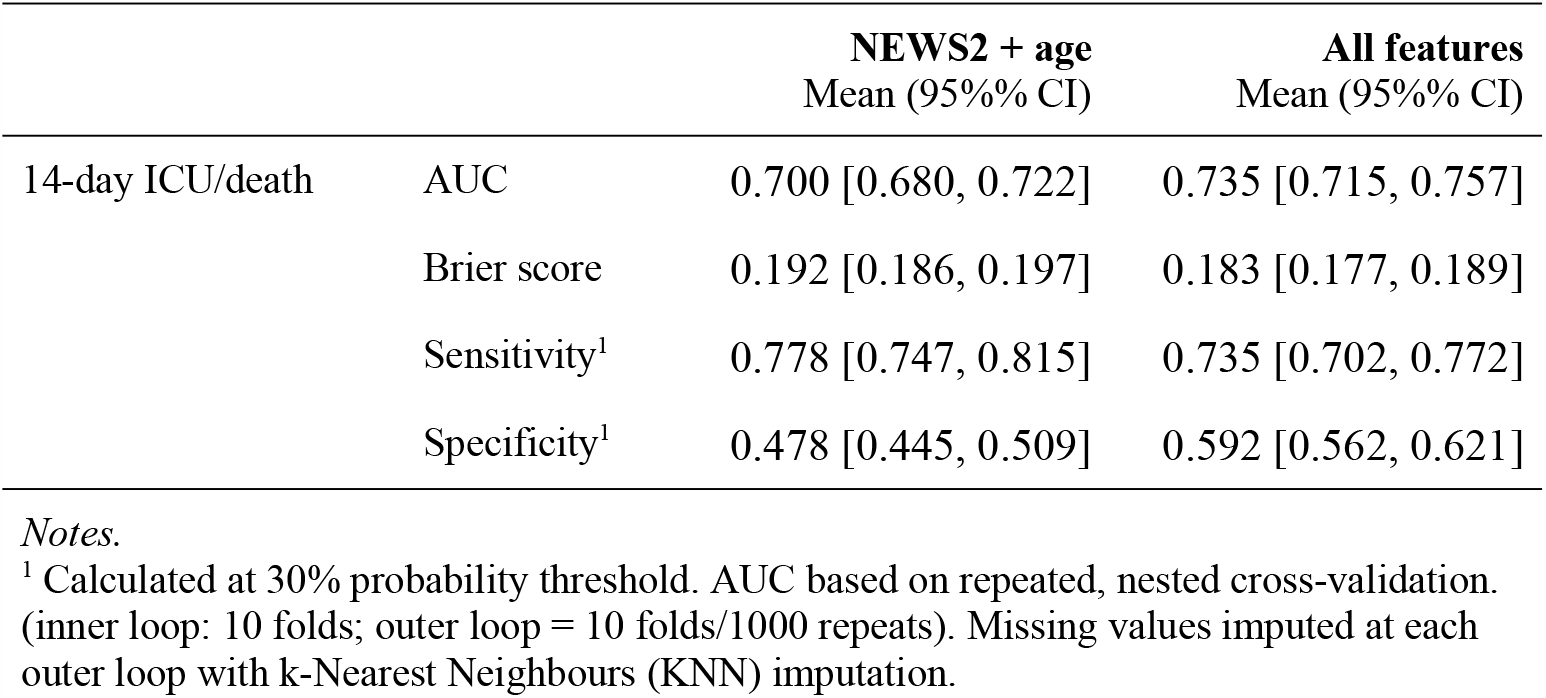
KCH internally validated predictive performance (n=1276) based on nested repeated cross-validation

|  |  | NEWS2 + age<br>Mean (95%% CI) | All features<br>Mean (95%% CI) |
| --- | --- | --- | --- |
| 14-day ICU/death | AUC | 0.700 [0.680, 0.722] | 0.735 [0.715, 0.757] |
|  | Brier score | 0.192 [0.186, 0.197] | 0.183 [0.177, 0.189] |
|  | Sensitivity<sup>1</sup> | 0.778 [0.747, 0.815] | 0.735 [0.702, 0.772] |
|  | Specificity<sup>1</sup> | 0.478 [0.445, 0.509] | 0.592 [0.562, 0.621] |

*Notes.*
<sup>1</sup> Calculated at 30% probability threshold. AUC based on repeated, nested cross-validation. (inner loop: 10 folds; outer loop = 10 folds/1000 repeats). Missing values imputed at each outer loop with k-Nearest Neighbours (KNN) imputation.

**Figure 1:**
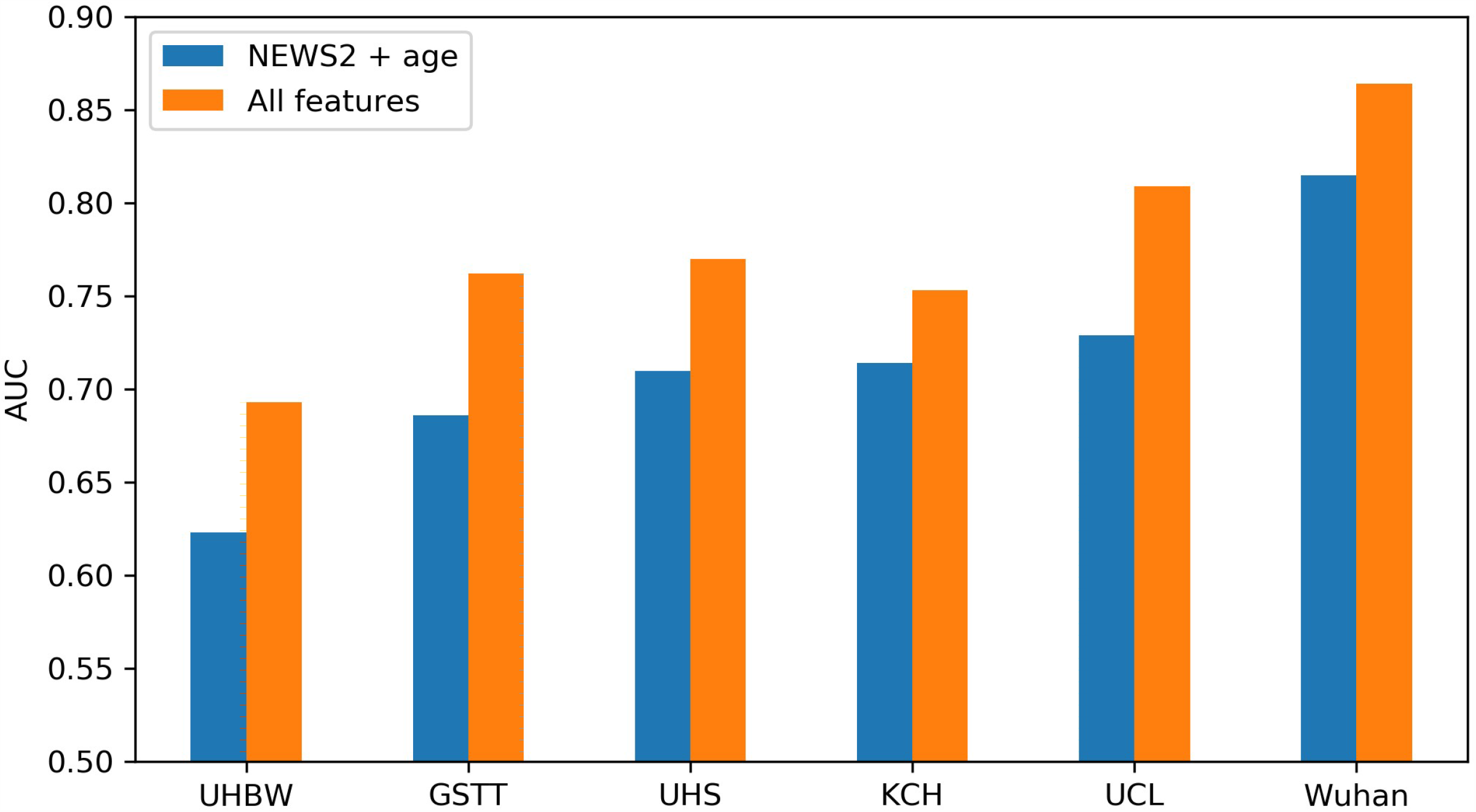
Improvement in discrimination comparing baseline model (‘NEWS2 + age’) with final model (‘All features’) for 14-day ICU transfer/death.

**Figure 2:**
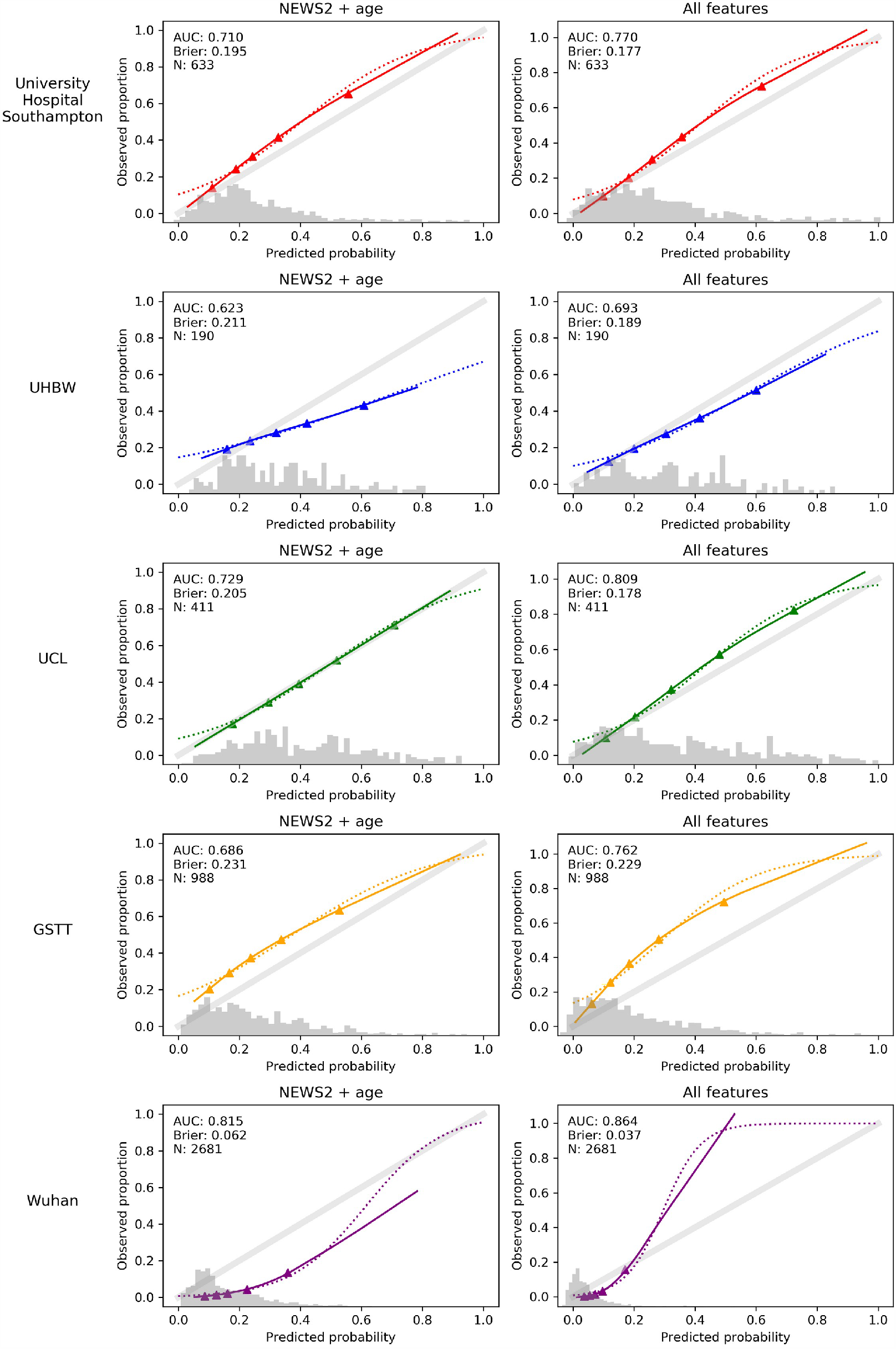
Discrimination and calibration curves for 14-day ICU/death at external validation.

#### Supplementing NEWS2 with routinely collected blood and physiological parameters

We considered whether routine blood and physiological parameters could improve risk stratification for medium-term COVID outcome (ICU transfer/death at 14 days). When adding demographic, blood, and physiological parameters to NEWS2, nine features were retained following LASSO regularisation, in order of effect size: NEWS2 score, supplemental oxygen flow rate, urea, age, oxygen saturation, CRP, estimated GFR, neutrophil count, neutrophil/lymphocyte ratio. Notably, comorbid conditions were not retained when added in subsequent models, suggesting most of the variance explained was already captured by the included parameters. Internally validated discrimination in the KCH training sample was moderate (AUC = 0.735; 95% CI: 0.715, 0.757) but improved compared to NEWS2 alone (Table 3). This improvement over NEWS2 alone was replicated in validation samples (Figure 1). The supplemented model continued to show evidence of substantial miscalibration.

### Sensitivity analyses

For the 3-day endpoint, 13% of patients at KCH (n=163) and between 17% and 29% of patients at UK NHS trusts were transferred to ICU or died (Table 1). The 3-day model retained just two parameters following regularisation: NEWS2 score and supplemental oxygen flow rate. For the baseline model (‘NEWS2 + age’) discrimination was moderate at internal validation (AUC = 0.764; 95% CI: 0.737, 0.794; Supplementary Table 3) and external validation (AUC = 0.657 to 0.755) but calibration remained poor (Supplementary Figure 1). Moreover, the supplemented model (‘NEWS2 + oxygen flow rate’) showed smaller improvements in discrimination compared to those seen at 14 days. For the KCH training cohort internally validated AUC increased by 0.025: from 0.764 (95% CI: 0.737, 0.794) for ‘NEWS2 + age’ to 0.789 (0.763, 0.819) for the supplemented model (‘NEWS2 + oxygen flow rate’). At external validation, improvements were modest (UHBW) or negative (GSTT) in some sites; but more substantial in others (UHS, UCL). Moreover, model calibration was considerably worse for the supplemented 3-day model (Supplementary Figure 1).

We found no evidence of difference by sex (results not shown) and findings were consistent when additionally adjusting for ethnicity in the subset of individuals with ethnicity data (Supplementary Tables 2 and 3); and when excluding nosocomial patients (Supplementary Tables 2 and 3).

### Decision curve analysis

Decision curve analysis for the 14-day endpoint is presented in Figure 3. At KCH the baseline model (‘NEWS2 + age’) offered small increments in net benefit compared to the ‘treat all’ and ‘treat none’ strategies for risk thresholds in the range 25% to 60%. This was replicated in all validation cohorts except for UHBW, where net benefit for ‘NEWS2 + age’ is lower than the ‘treat none’ strategy beyond the 40% risk threshold. The supplemented model (‘All features’) improves upon ‘NEWS2 + age’ and the two default strategies in most sites across the range 20% to 80%, except for (i) UHBW, where ‘treat none’ is superior beyond a threshold of 55%; (ii) GSTT, where ‘treat all’ is superior up to a threshold of 30%, and there is no improvement for supplemented model.

**Figure 3:**
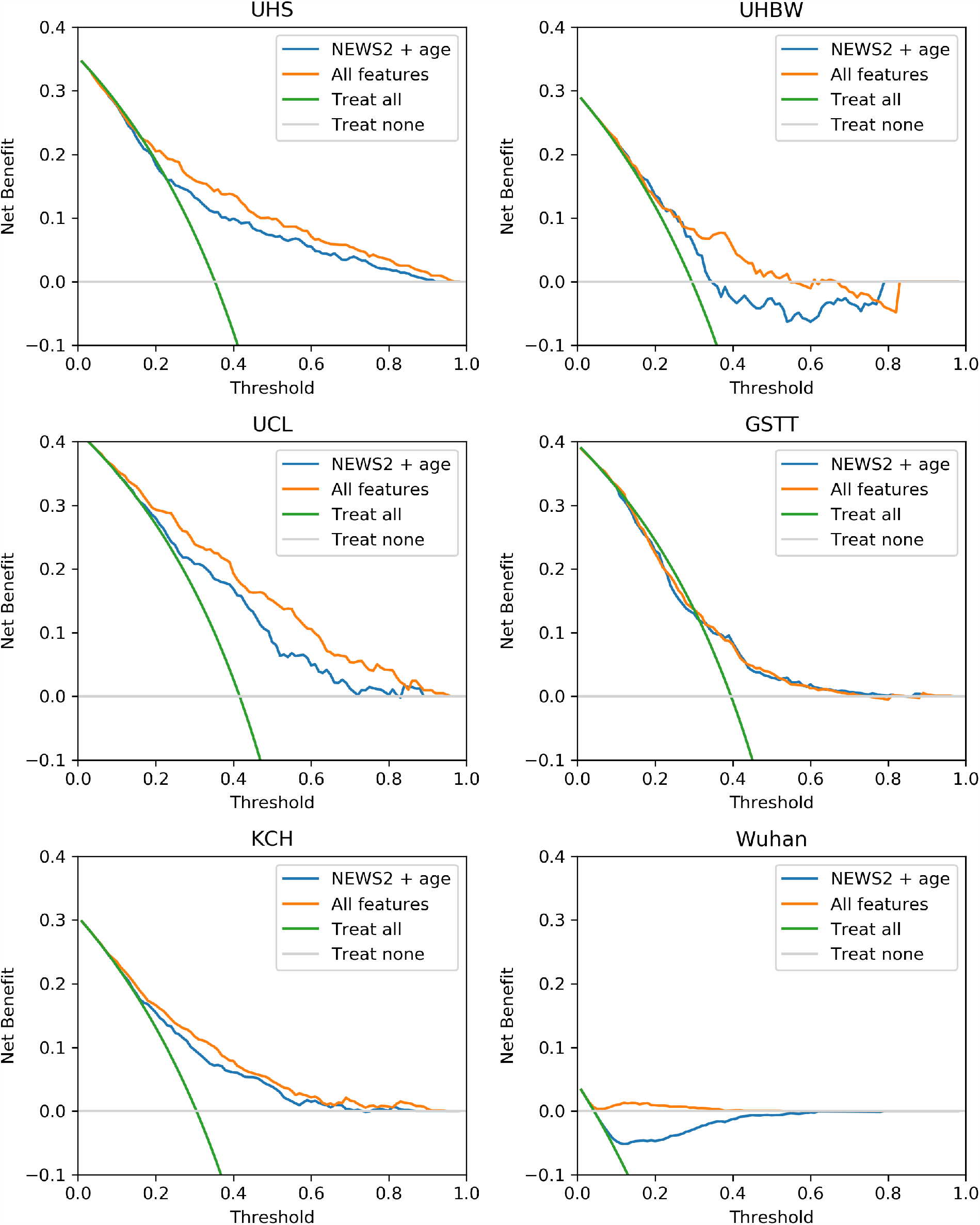
Net benefit for baseline and supplemented model at 14-day endpoint, compared with ‘Treat all’ and ‘Treat none’ default strategies.

For the 3-day endpoint the improvement in net benefit for the supplemented model over the two default strategies was smaller, compared to improvements seen at 14 days (Supplementary Figure 2). At two sites (UHBW and GSST) neither the baseline (‘NEWS2 + age’) supplemented (‘All features’) model offered any improvement over the ‘treat all’ or ‘treat none’ strategies. At KCH and UHS net benefit for ‘NEWS2 + age’ was higher than the default strategies for a range of risk thresholds, but was not increased further by the supplemented (‘NEWS2 + oxygen flow rate’) model.

## Discussion

### Principal findings

To our knowledge, this is the first study to systematically evaluate the UK NEWS2 acuity score for severe COVID-19 outcome, and the first to externally evaluate it beyond national sites (four UK NHS Trusts and two hospitals in Wuhan, China). We found that while ‘NEWS2 + age’ had moderate discrimination for short-term COVID outcome (3-day ICU transfer/death), it showed poor-to-moderate discrimination for medium-term outcome (14-day ICU transfer/death), questioning its suitability as a screening tool for COVID patients. Risk stratification was improved by adding routinely-collected blood and physiological parameters, and discrimination in supplemented models was moderate-to-good. However, the model showed evidence of miscalibration, with a tendency to underestimate risks in external sites. The derived model for 14-day ICU transfer/death included nine parameters: NEWS2 score, supplemental oxygen flow rate, urea, age, oxygen saturation, CRP, estimated GFR, neutrophil count, neutrophil/lymphocyte ratio. Notably, pre-existing comorbidities did not improve risk prediction and were not retained in the final model. This was unexpected but may indicate that the effect of pre-existing health conditions could be manifest through some of the included blood or physiological markers.

Overall, this study overcomes many of the factors associated with high risk of bias in the development of prognostic models for COVID-19(13) and provides some evidence to support the supplementation of NEWS2 for clinical decisions with these patients.

### Comparison with other studies

A systematic review of 10 prediction models for mortality in COVID-19 infection(10) found broad similarities with the features retained in our models, particularly regarding CRP and neutrophil levels. However, existing prediction models suffer several methodological weaknesses including over-fitting, selection bias, and reliance on cross-sectional data without accounting for censoring. Additionally, many existing studies have relied on single centre studies or in ethnically homogenous Chinese cohorts, whereas the present study shows validation in multiple organisations and diverse populations. A key strength of our study is the robust and repeated external validation across national and international sites; however evidence of miscalibration suggests we should be cautious when attempting to generalise these findings. Future research should include larger collaborations and aim to develop ‘from onset’ population predictions.

NEWS2 is a summary score derived from six physiological parameters, including oxygen supplementation. Lack of evidence for NEWS2 use in COVID-19 especially in primary care has been highlighted(9). The oxygen saturation component of physiological measurements added value beyond NEWS2 total score and was retained following regularisation for 14-day endpoints. This suggests some residual association over and above what is captured by the NEWS2 score, and reinforces Royal College of Physicians guidance that the NEWS2 score ceilings with respect to respiratory function(35).

Cardiac disease and myocardial injury have been described in severe COVID-19 cases in China(2,23). In our model, blood Troponin-T, a marker of myocardial injury, had additional salient signal but was only measured in a subset of our cohort at admission, so it was excluded from our final model. This could be explored further in larger datasets.

### Strengths and limitations

Our study provides a risk stratification model for which we obtained generalisable and robust results across UK and non-UK sites with differing geographical catchment and population characteristics. However, some limitations must be acknowledged. First, there are likely to be other parameters not measured in this study that could substantially improve the risk stratification model (e.g. radiological features or comorbidity load). These parameters could be explored in future work but were not considered in the present study to avoid limiting the real-world implementation of the risk stratification model. Second, our models showed better performance in UK secondary care settings among populations with higher rates of severe COVID disease. Therefore, further research is needed to investigate the suitability of our model for primary care settings which have a high prevalence of mild disease severities and in community settings. This would allow us to capture variability at earlier stages of the disease and trends in patients not requiring hospital admission. Third, while external validation across multiple national and international sites represents a key strength, we did not have access to individual participant data and model development was limited to a single site (i.e. KCH). Although we benefited from existing infrastructure to support rapid data analysis, we urgently need infrastructure to support data sharing between sites to address some of the limitations of the present study (e.g. miscalibration) and improve the transferability of these models. This would facilitate not only external validation but, more importantly, would make it possible for prediction models to be developed across sites using pooled, individual participant data(36).

## Conclusions

The NEWS2 early warning score is in near-universal use in UK NHS Trusts since March 2019(15) but little is known about its use for COVID patients. Here we showed that NEWS2 and age at hospital admission had moderate discrimination for medium-term (14-day) severe COVID outcome, questioning its use as a tool to guide hospital admission. Moreover, we showed that NEWS2 discrimination could be improved by adding eight blood and physiological parameters (supplemental oxygen flow rate, urea, age, oxygen saturation, CRP, estimated GFR, neutrophil count, neutrophil/lymphocyte ratio) which are routinely collected and readily available in healthcare services. Thus, this type of model could be easily implemented in clinical practice and predicted risk score probabilities of individual patients are easy to communicate. At the same time, although we provided some evidence of improved discrimination versus NEWS2 and age alone, given miscalibration in external sites, our proposed model should be used as a complement and not as a replacement for clinical judgment.

## Data Availability

Code and pre-trained models are available at https://github.com/ewancarr/NEWS2-COVID-19 and openly shared for testing in other COVID datasets.

Source text from patient records used at all sites in the study will not be available due to inability to safely fully anonymise up to the Information Commissioner Office (ICO) standards and would be likely to contain strong identifiers (e.g. names, postcodes) and highly sensitive data (e.g. diagnoses).

A subset of the KCH dataset limited to anonymisable information (e.g. only SNOMED codes and aggregated demographics) is available on request to researchers with suitable training in information governance and human confidentiality protocols subject to approval by the King's College Hospital Information Governance committee; applications for research access should be sent to. This dataset cannot be released publicly due to the risk of re-identification of such granular individual level data, as determined by the King's College Hospital Caldicott Guardian.

The GSTT dataset cannot be released publicly due to the risk of re-identification of such granular individual level data, as determined by the Guy's and St. Thomas's Trust Caldicott Guardian.

The UHS dataset cannot be released publicly due to the risk of re-identification of such granular individual level data, as determined by the University Hospital Southampton Caldicott Guardian.

The Wuhan dataset used in the study will not be available due to inability to fully anonymise in line with ethical requirements. Applications for research access should be sent to TS and details will be made available via https://covid.datahelps.life/prediction/.

https://github.com/ewancarr/NEWS2-COVID-19

## Declarations

### Ethics approval and consent to participate

The KCH component of the project operated under London South East Research Ethics Committee (reference 18/LO/2048) approval granted to the King’s Electronic Records Research Interface (KERRI); specific work on COVID-19 research was reviewed with expert patient input on a virtual committee with Caldicott Guardian oversight. The UHS validation was performed as part of a service evaluation agreed with approval from trust research leads and the Caldicott Guardian. Ethical approval for GSTT was granted by The London Bromley Research Ethics Committee (reference 20/HRA/1871) to the King’s Health Partners Data Analytics and Modelling COVID-19 Group to collect clinically relevant data points from patient’s electronic health records. The Wuhan validation was approved by the Research Ethics Committee of Shanghai Dongfang Hospital and Taikang Tongji Hospital.

### Consent for publication

Not applicable.

### Availability of data and materials

The UCH data cannot be released publicly due to conditions of regulatory approvals that preclude open access data sharing to minimise risk of patient identification through granular individual health record data. The authors will consider specific requests for data sharing as part of academic collaborations subject to ethical approval and data transfer agreements in accordance with GDPR regulations.

## Competing interests

JTHT received research support and funding from InnovateUK, Bristol-Myers-Squibb, iRhythm Technologies, and holds shares <£5,000 in Glaxo Smithkline and Biogen.

## Funding and Acknowledgments

DMB is funded by a UKRI Innovation Fellowship as part of Health Data Research UK MR/S00310X/1 (https://www.hdruk.ac.uk).

RB is funded in part by grant MR/R016372/1 for the King’s College London MRC Skills Development Fellowship programme funded by the UK Medical Research Council (MRC, https://mrc.ukri.org) and by grant IS-BRC-1215-20018 for the National Institute for Health Research (NIHR, https://www.nihr.ac.uk) Biomedical Research Centre at South London and Maudsley NHS Foundation Trust and King’s College London.

RJBD is supported by: (1) NIHR Biomedical Research Centre at South London and Maudsley NHS Foundation Trust and King’s College London, London, U.K. (2) Health Data Research UK, which is funded by the UK Medical Research Council, Engineering and Physical Sciences Research Council, Economic and Social Research Council, Department of Health and Social Care (England), Chief Scientist Office of the Scottish Government Health and Social Care Directorates, Health and Social Care Research and Development Division (Welsh Government), Public Health Agency (Northern Ireland), British Heart Foundation and Wellcome Trust. (3) The BigData@Heart Consortium, funded by the Innovative Medicines Initiative-2 Joint Undertaking under grant agreement No. 116074. This Joint Undertaking receives support from the European Union’s Horizon 2020 research and innovation programme and EFPIA; it is chaired by DE Grobbee and SD Anker, partnering with 20 academic and industry partners and ESC. (4) The National Institute for Health Research University College London Hospitals Biomedical Research Centre. (5) National Institute for Health Research (NIHR) Biomedical Research Centre at South London and Maudsley NHS Foundation Trust and King’s College London. (5) The UK Research and Innovation London Medical Imaging & Artificial Intelligence Centre for Value Based Healthcare (6) the National Institute for Health Research (NIHR) Applied Research Collaboration South London (NIHR ARC South London) at King’s College Hospital NHS Foundation Trust.

KO’G is supported by an MRC Clinical Training Fellowship (MR/R017751/1).

WW is supported by a Health Foundation grant.

AD and VC acknowledge support from National Institute for Health Research (NIHR) Applied Research Collaboration (ARC) South London at King’s College Hospital NHS Foundation Trust and the Royal College of Physicians, as well as the support from the NIHR Biomedical Research Centre based at Guy’s and St Thomas’ NHS Foundation Trust and King’s College London. VC is additionally supported by Health Data Research UK, which is funded by the UK Medical Research Council, Engineering and Physical Sciences Research Council, Economic and Social Research Council, Department of Health and Social Care (England), Chief Scientist Office of the Scottish Government Health and Social Care Directorates, Health and Social Care Research and Development Division (Welsh Government), Public Health Agency (Northern Ireland), British Heart Foundation and Wellcome Trust.

RZ is supported by a King’s Prize Fellowship.

AS is supported by a King’s Medical Research Trust studentship.

JTHT is supported by London AI Medical Imaging Centre for Value-Based Healthcare (AI4VBH) and the National Institute for Health Research (NIHR) Applied Research Collaboration South London (NIHR ARC South London) at King’s College Hospital NHS Foundation Trust.

FB and PTTH are funded by National Institute for Health Research (NIHR) Biomedical Research Centre, Data Sciences at University Hospital Southampton NHS Foundation Trust and the Clinical Informatics Research Unit, University of Southampton.

JB is funded by the Clinical Informatics Research Unit, University of Southampton, and part funded by the Global Alliance for Chronic Disease (GDAC).

AP is part funded by UHS Digital, University Hospital Southampton, Tremona Road, Southampton.

AJS is supported by a Digital Health Fellowship through Health Education England (Wessex).

HW and HZ are supported by Medical Research Council and Health Data Research UK Grant (MR/S004149/1), Industrial Strategy Challenge Grant (MC_PC_18029) and Wellcome Institutional Translation Partnership Award (PIII054). XW is supported by National Natural Science Foundation of China (grant number 81700006).

AMS is supported by the British Heart Foundation (CH/1999001/11735), the National Institute for Health Research (NIHR) Biomedical Research Centre at Guy’s & St Thomas’ NHS Foundation Trust and King’s College London (IS-BRC-1215-20006), and the Fondation Leducq. AP is partially supported by NIHR NF-SI-0617-10120. This work was supported by the National Institute for Health Research (NIHR) University College London Hospitals (UCH) Biomedical

Research Centre (BRC) Clinical and Research Informatics Unit (CRIU), NIHR Health Informatics Collaborative (HIC), and by awards establishing the Institute of Health Informatics at University College London (UCL). This work was also supported by Health Data Research UK, which is funded by the UK Medical Research Council, Engineering and Physical Sciences Research Council, Economic and Social Research Council, Department of Health and Social Care (England), Chief Scientist Office of the Scottish Government Health and Social Care Directorates, Health and Social Care Research and Development Division (Welsh Government), Public Health Agency (Northern Ireland), British Heart Foundation and the Wellcome Trust.

RKG is funded by the NIHR (DRF-2018-11-ST2-004); MN is funded by the Wellcome Trust (207511/Z/17/Z).

This paper represents independent research part-funded by the National Institute for Health Research (NIHR) Biomedical Research Centres at South London and Maudsley NHS Foundation Trust, London AI Medical Imaging Centre for Value-Based Healthcare, and Guy’s & St Thomas’ NHS Foundation Trust, both with King’s College London. The views expressed are those of the author(s) and not necessarily those of the NHS, the NIHR or the Department of Health and Social Care. The funders had no role in study design, data collection and analysis, decision to publish, or preparation of the manuscript. We would also like to thank all the clinicians managing the patients, the patient experts of the KERRI committee, Professor Irene Higginson, Professor Alastair Baker, Professor Jules Wendon, Dan Persson and Damian Lewsley for their support.

The authors acknowledge use of the research computing facility at King’s College London, Rosalind (https://rosalind.kcl.ac.uk), which is delivered in partnership with the National Institute for Health Research (NIHR) Biomedical Research Centres at South London & Maudsley and Guy’s & St. Thomas’ NHS Foundation Trusts, and part-funded by capital equipment grants from the Maudsley Charity (award 980) and Guy’s & St. Thomas’ Charity (TR130505). The views expressed are those of the author(s) and not necessarily those of the NHS, the NIHR, King’s College London, or the Department of Health and Social Care.

## Authors’ contributions

The corresponding author, Dr Ewan Carr, is guarantor of the manuscript.

JT, AS, RD, EC and RB conceived the study design and developed the study objectives. JT, RD, AF, LR, DB, ZK, TS and AS were the leads to develop CogStack platform. DB, ZK, TS, AS were responsible for the data extraction and preparation. EC, RB, AP, DS contributed to the statistical analyses. All authors contributed to the interpretation of the data. AS, JT, KO, RZ provided clinical input. All authors contributed to interpret the data, draft the article and provided final approval of the manuscript. DMB, ZK, AS, TS, JTHT, LR, KN performed data processing and software development; KOG, RZ, JTHT performed data validation.

At GSTT, WW and WM were responsible for the data extraction and preparation. WW performed the model validation. AD and VC contributed to the interpretation of the data.

At UHS, MS and FB were responsible for the data extraction and preparation. MS, HP and AS contributed to the statistical analysis. All authors contributed to the interpretation of the data. MS and AP provided clinical input. MS and HP performed data/model validation.

For the Wuhan cohort, XZ, XW and JS extracted the data from the EHR system. HW and HZ preprocessed the raw data and conducted the prediction model validations. BG, HW, HZ, TS and JS interpreted the data and results.

The views expressed are those of the authors and not necessarily those of the MRC, NHS, the NIHR or the Department of Health and Social Care. The funders of the study had no role in the study design, data collection, data analysis, data interpretation, writing of the report or the decision to submit the article for publication.

## Supplementary Methods

### Data Processing

#### King’s College Hospital

Data was extracted from the structured and unstructured components of the electronic health record (EHR) using natural language processing (NLP) tools belonging to the CogStack ecosystem(37), namely MedCAT(38) and MedCATTrainer(39). The CogStack NLP pipeline captures negation, synonyms, and acronyms for medical SNOMED-CT concepts as well as surrounding linguistic context using deep learning and long short-term memory networks. MedCAT produces unsupervised annotations for all SNOMED-CT concepts (Supplementary Table 4) under parent terms Clinical Finding, Disorder, Organism, and Event with disambiguation, pre-trained on MIMIC-III(40).

Starting from our previous model(41), further supervised training improved detection of annotations and meta-annotations such as experiencer (is the concept annotated experienced by the patient or other), negation (is the concept annotated negated or not) and temporality (is the concept annotated in the past or present) with MedCATTrainer. Meta-annotations for hypothetical, historical and experiencer were merged into “Irrelevant” allowing us to exclude any mentions of a concept that do not directly relate to the patient currently. Performance of the NLP pipeline for comorbidities mentioned in the text was evaluated on 4343 annotations in 146 clinical documents by a clinician (JT). F1 scores, precision, and recall are presented in Supplementary Table 5.

##### Guy’s and St Thomas NHS Foundation Trust (GSTT)

Electronic health records from all patients admitted to Guy’s and St Thomas NHS Foundation Trust who had a positive COVID-19 test result between the 3^rd^ of March and 21^st^ of May 2020, inclusive, were identified. Data were extracted using structured queries from six complementary platforms and linked using unique patient identifiers. Data processing was performed using Python 3.7(30). The process and outputs were reviewed by a study clinician.

##### University Hospitals Southampton (UHS)

Data were extracted from the structured components of the UHS CHARTS EHR system and data warehouse. Data was transformed to the required format for validation purposes using Python 3.7(30). Diagnosis and comorbidity data of interest were gathered from ICD-10 coded data. No unstructured data extraction was required for validation purposes. The process and outputs were reviewed by an experienced clinician prior to analysis.

##### University College Hospital London (UCH)

Dates of hospital admission, symptom onset, ICU transfer and death were extracted from electronic health records. The outcome (14 day ICU/death) was defined in UCLH as “initiation of ventilatory support (continuous positive airway pressure, non-invasive ventilation, high flow nasal cannula oxygen, invasive mechanical ventilation or extracorporeal membrane oxygenation) or death” which is consistent WHO-COVID-19 Outcomes Scales 6-8.

##### Wuhan cohort

Demographic, premorbid conditions, clinical symptoms or signs at presentation, laboratory data, treatment and outcome data were extracted from electronic medical records using a standardised data collection form by a team of experienced respiratory clinicians, with double data checking and involvement of a third reviewer where there was disagreement. Anonymised data was entered into a password-protected computerised database.

---

**Supplementary Figure 1:**
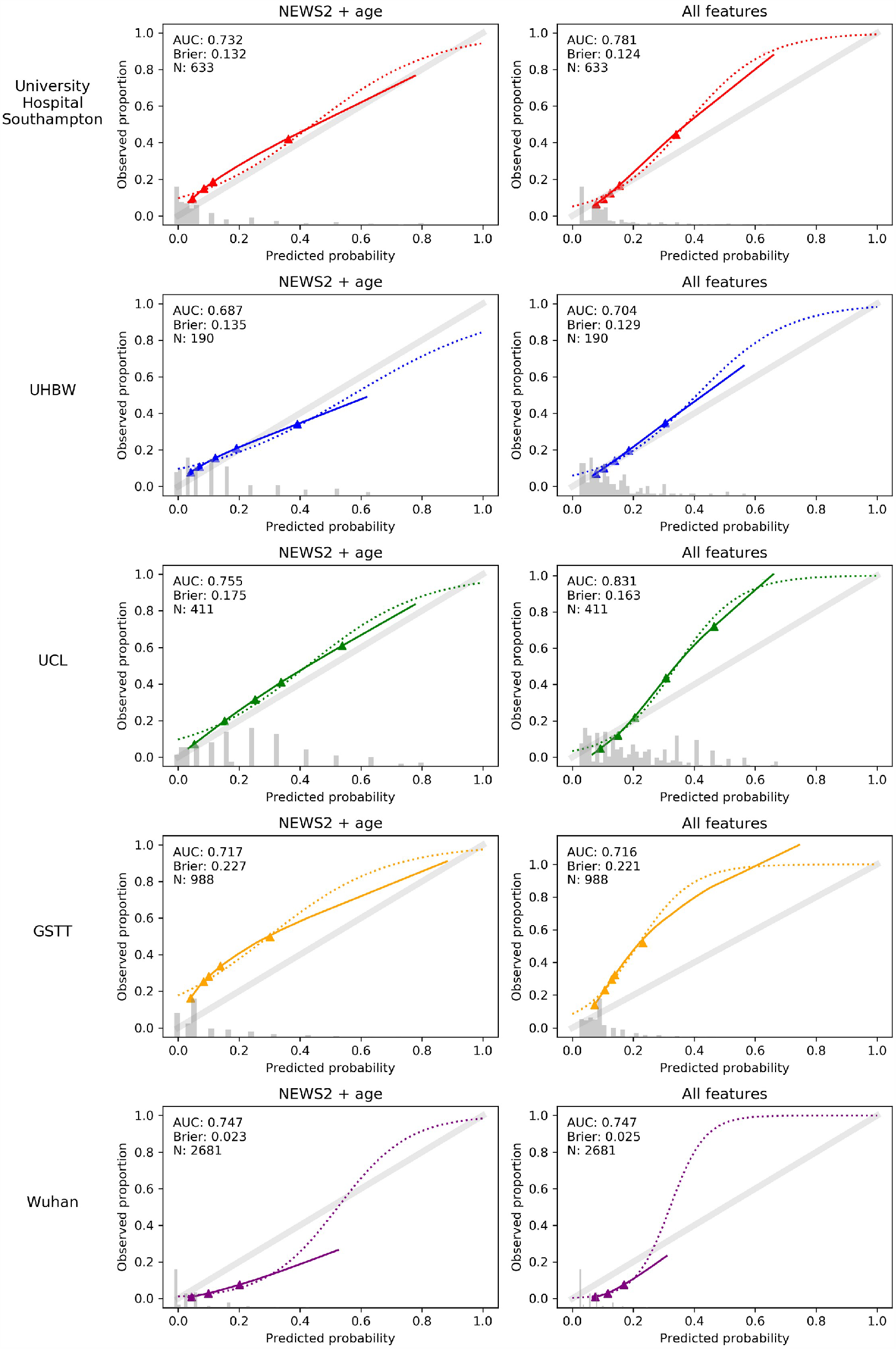
Discrimination and calibration curves for 3-day ICU/death at external validation.

**Supplementary Figure 2:**
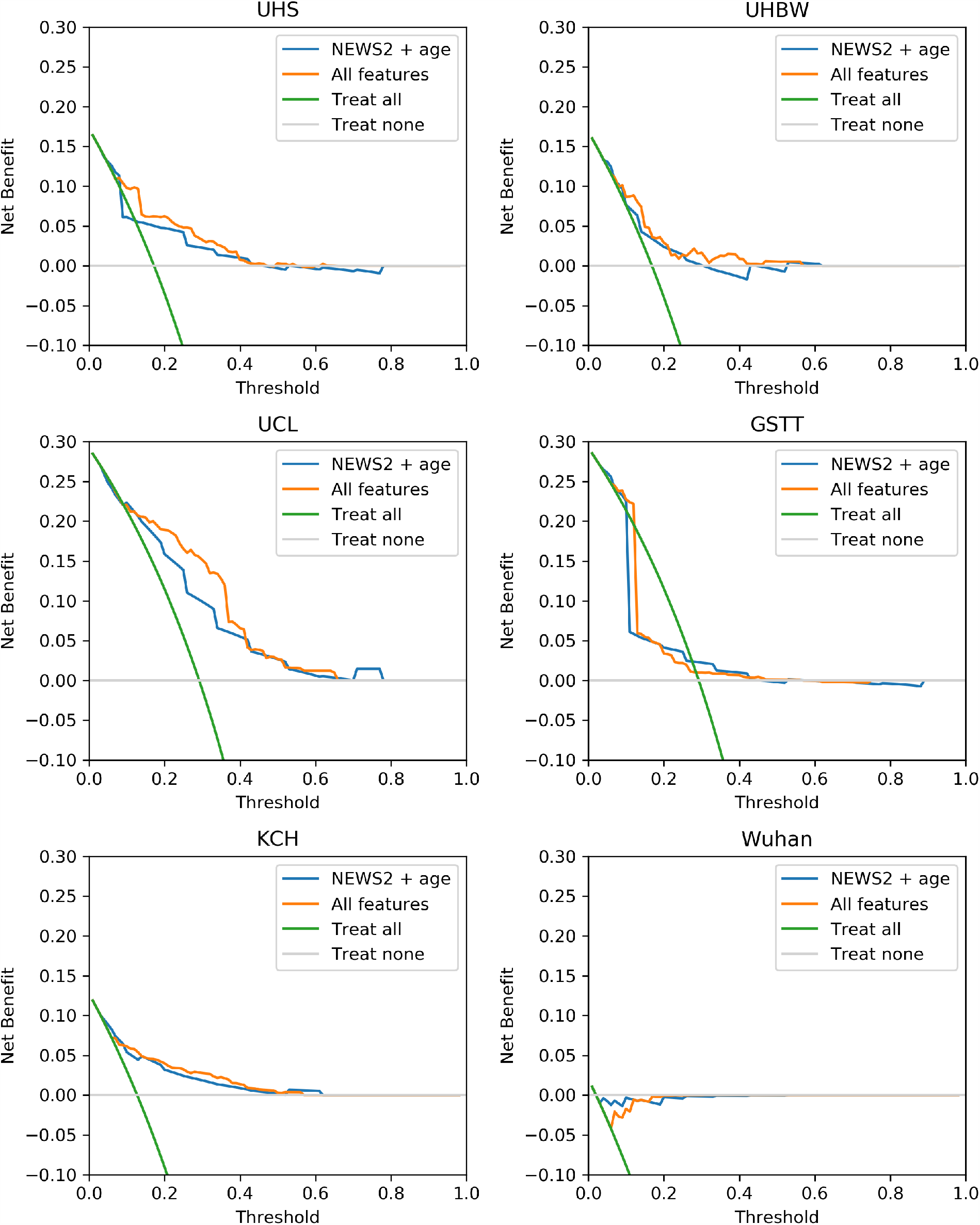
Net benefit for baseline and supplemented model at 3-day endpoint, compared with ‘Treat all’ and ‘Treat none’ default strategies.

**Supplementary Table 1:** Logistic regression models for each blood and physiological measure tested separately in the KCH training cohort, for 14- and 3-day ICU/death

|  |  | 14 day ICU/death |  |  |  | 3 day ICU/death |  |  |  |
| --- | --- | --- | --- | --- | --- | --- | --- | --- | --- |
|  |  | Model 1: Age, sex only<sup>1</sup> |  | Model 2: + comorbidities<sup>2</sup> |  | Model 1: Age, sex only<sup>1</sup> |  | Model 2: + comorbidities<sup>2</sup> |  |
|  | N avail. | Odds Ratio<br>[95% C.I.] | *P*-value<sup>3</sup> | Odds Ratio<br>[95% C.I.] | *P*-value<sup>3</sup> | Odds Ratio<br>[95% C.I.] | *P*-value<sup>3</sup> | Odds Ratio<br>[95% C.I.] | *P*-value<sup>3</sup> |
| NEWS2 score | 1262 | 2.10 [1.83, 2.40] | <0.001 | 2.13 [1.86, 2.44] | <0.001 | 2.66 [2.25, 3.16] | <0.001 | 2.65 [2.23, 3.15] | <0.001 |
| Oxygen flow rate | 1271 | 1.92 [1.70, 2.17] | <0.001 | 1.97 [1.74, 2.22] | <0.001 | 2.22 [1.92, 2.56] | <0.001 | 2.25 [1.94, 2.60] | <0.001 |
| Respiratory rate | 1273 | 1.65 [1.45, 1.86] | <0.001 | 1.65 [1.45, 1.87] | <0.001 | 1.83 [1.59, 2.10] | <0.001 | 1.81 [1.58, 2.09] | <0.001 |
| CRP | 1240 | 1.64 [1.44, 1.87] | <0.001 | 1.65 [1.45, 1.88] | <0.001 | 1.89 [1.59, 2.24] | <0.001 | 1.92 [1.61, 2.28] | <0.001 |
| Lymphocyte/CRP ratio | 1196 | 0.63 [0.55, 0.73] | <0.001 | 0.63 [0.54, 0.72] | <0.001 | 0.58 [0.47, 0.71] | <0.001 | 0.57 [0.47, 0.71] | <0.001 |
| Oxygen saturation | 1273 | 0.65 [0.57, 0.73] | <0.001 | 0.63 [0.56, 0.72] | <0.001 | 0.51 [0.44, 0.60] | <0.001 | 0.49 [0.42, 0.58] | <0.001 |
| Neutrophil count | 1220 | 1.41 [1.25, 1.60] | <0.001 | 1.42 [1.25, 1.61] | <0.001 | 1.53 [1.31, 1.80] | <0.001 | 1.54 [1.31, 1.81] | <0.001 |
| Urea | 1221 | 1.40 [1.23, 1.60] | <0.001 | 1.42 [1.23, 1.64] | <0.001 | 1.38 [1.17, 1.63] | <0.001 | 1.48 [1.23, 1.78] | <0.001 |
| Neutrophil/lymphocyte ratio | 1218 | 1.38 [1.21, 1.57] | <0.001 | 1.39 [1.21, 1.58] | <0.001 | 1.44 [1.21, 1.73] | <0.001 | 1.45 [1.21, 1.74] | <0.001 |
| Heart rate | 1273 | 1.34 [1.18, 1.52] | <0.001 | 1.35 [1.19, 1.53] | <0.001 | 1.54 [1.30, 1.82] | <0.001 | 1.54 [1.30, 1.82] | <0.001 |
| Estimated GFR | 1254 | 0.78 [0.68, 0.88] | <0.001 | 0.76 [0.66, 0.88] | <0.001 | 0.87 [0.73, 1.03] | 0.125 | 0.82 [0.67, 1.00] | 0.058 |
| Albumin | 1153 | 0.80 [0.70, 0.92] | 0.002 | 0.81 [0.71, 0.92] | 0.002 | 0.76 [0.64, 0.90] | 0.002 | 0.75 [0.63, 0.89] | 0.002 |
| Temperature | 1273 | 1.21 [1.07, 1.37] | 0.004 | 1.21 [1.07, 1.38] | 0.003 | 1.23 [1.04, 1.44] | 0.018 | 1.23 [1.05, 1.45] | 0.016 |
| Platelet count | 1224 | 0.88 [0.78, 1.01] | 0.075 | 0.88 [0.78, 1.01] | 0.080 | 1.05 [0.89, 1.24] | 0.630 | 1.04 [0.88, 1.23] | 0.698 |
| Diastolic blood pressure | 1273 | 0.89 [0.78, 1.00] | 0.075 | 0.89 [0.78, 1.01] | 0.080 | 0.80 [0.67, 0.95] | 0.016 | 0.79 [0.66, 0.94] | 0.012 |
| Systolic blood pressure | 1273 | 0.91 [0.81, 1.03] | 0.157 | 0.91 [0.80, 1.03] | 0.148 | 0.95 [0.80, 1.13] | 0.625 | 0.94 [0.79, 1.12] | 0.522 |
| Lymphocytes | 1221 | 0.93 [0.82, 1.06] | 0.279 | 0.93 [0.82, 1.06] | 0.282 | 1.01 [0.85, 1.19] | 0.942 | 1.00 [0.84, 1.19] | 0.980 |
| Hemoglobin | 1223 | 1.04 [0.92, 1.19] | 0.505 | 1.07 [0.94, 1.22] | 0.329 | 1.20 [1.01, 1.44] | 0.052 | 1.21 [1.00, 1.45] | 0.057 |

*Notes.*
Continuous predictors were standardised (mean = 0; standard deviation = 1), therefore, odds ratios represent one standard deviation change in the respective parameter, each tested in a separate model. <sup>1</sup>Adjusted for age and sex only. <sup>2</sup>Additionally adjusted for comorbidities (hypertension, diabetes, heart diseases, respiratory diseases and chronic kidney disease). <sup>3</sup>FDR-corrected *P*-values based on the Benjamini-Hochberg correction.

**Supplementary Table 2:** Univariate logistic regression models for sensitivity analyses

|  |  |  |  |  |  |  |  |  |  |  |  |  |
| --- | --- | --- | --- | --- | --- | --- | --- | --- | --- | --- | --- | --- |
| 14-day ICU/death |  |  |  |  |  |  | 3-day ICU/death |  |  |  |  |  |
|  | Subset 1: Adjusted for ethnicity, in the subset of patients with non-missing ethnicity |  |  | Subset 2: Excluding nosocomial patients |  |  | Subset 1: Adjusted for ethnicity, in the subset of patients with non-missing ethnicity |  |  | Subset 2: Excluding nosocomial patients |  |  |
|  | N | OR [95% CI] | P-value | N | OR [95% CI] | P-value | N | OR [95% CI] | P-value | N | OR [95% CI] | P-value |
| NEWS2 score | 948 | 2.07 [1.78, 2.41] | <0.001 | 1109 | 2.27 [1.96, 2.63] | <0.001 | 948 | 2.50 [2.06, 3.03] | <0.001 | 1109 | 2.75 [2.29, 3.31] | <0.001 |
| Oxygen flow rate | 955 | 1.80 [1.57, 2.07] | <0.001 | 1118 | 2.01 [1.77, 2.29] | <0.001 | 955 | 2.12 [1.80, 2.50] | <0.001 | 1118 | 2.30 [1.97, 2.67] | <0.001 |
| Oxygen saturation | 957 | 0.60 [0.51, 0.69] | <0.001 | 1120 | 0.63 [0.55, 0.72] | <0.001 | 957 | 0.47 [0.39, 0.57] | <0.001 | 1120 | 0.51 [0.43, 0.60] | <0.001 |
| CRP | 932 | 1.71 [1.47, 1.99] | <0.001 | 1092 | 1.61 [1.40, 1.85] | <0.001 | 932 | 1.97 [1.60, 2.42] | <0.001 | 1092 | 1.86 [1.55, 2.23] | <0.001 |
| Respiratory rate | 957 | 1.61 [1.40, 1.85] | <0.001 | 1120 | 1.66 [1.46, 1.89] | <0.001 | 957 | 1.68 [1.43, 1.96] | <0.001 | 1120 | 1.85 [1.60, 2.14] | <0.001 |
| Lymphocyte/CRP ratio | 900 | 0.59 [0.50, 0.71] | <0.001 | 1056 | 0.64 [0.55, 0.75] | <0.001 | 900 | 0.58 [0.46, 0.74] | <0.001 | 1056 | 0.60 [0.48, 0.74] | <0.001 |
| Heart rate | 957 | 1.36 [1.18, 1.56] | <0.001 | 1120 | 1.39 [1.22, 1.59] | <0.001 | 957 | 1.59 [1.31, 1.92] | <0.001 | 1120 | 1.53 [1.29, 1.83] | <0.001 |
| Neutrophil count | 918 | 1.49 [1.29, 1.73] | <0.001 | 1075 | 1.34 [1.17, 1.54] | <0.001 | 918 | 1.63 [1.35, 1.97] | <0.001 | 1075 | 1.55 [1.30, 1.85] | <0.001 |
| Neutrophil/lymphocyte ratio | 917 | 1.47 [1.26, 1.71] | <0.001 | 1074 | 1.33 [1.15, 1.53] | <0.001 | 917 | 1.50 [1.21, 1.85] | <0.001 | 1074 | 1.41 [1.16, 1.71] | <0.001 |
| Urea | 917 | 1.38 [1.17, 1.62] | <0.001 | 1084 | 1.36 [1.17, 1.58] | <0.001 | 917 | 1.41 [1.15, 1.74] | 0.002 | 1084 | 1.43 [1.18, 1.73] | <0.001 |
| Albumin | 866 | 0.81 [0.69, 0.94] | 0.008 | 1036 | 0.80 [0.68, 0.93] | 0.005 | 866 | 0.75 [0.61, 0.92] | 0.009 | 1036 | 0.69 [0.56, 0.84] | <0.001 |
| Diastolic blood pressure | 957 | 0.91 [0.79, 1.05] | 0.22 | 1120 | 0.88 [0.77, 1.00] | 0.071 | 957 | 0.80 [0.65, 0.99] | 0.056 | 1120 | 0.77 [0.64, 0.93] | 0.01 |
| Temperature | 957 | 1.22 [1.06, 1.41] | 0.008 | 1120 | 1.28 [1.12, 1.46] | <0.001 | 957 | 1.19 [0.99, 1.43] | 0.082 | 1120 | 1.22 [1.03, 1.44] | 0.025 |
| Hemoglobin | 921 | 1.07 [0.92, 1.25] | 0.374 | 1079 | 1.11 [0.95, 1.28] | 0.215 | 921 | 1.23 [1.00, 1.52] | 0.071 | 1079 | 1.19 [0.97, 1.45] | 0.116 |
| Estimated GFR | 944 | 0.80 [0.69, 0.93] | 0.003 | 1108 | 0.78 [0.68, 0.90] | 0.005 | 944 | 0.92 [0.75, 1.12] | 0.137 | 1108 | 0.87 [0.72, 1.04] | 0.116 |
| Systolic blood pressure | 957 | 0.90 [0.78, 1.04] | 0.177 | 1120 | 0.93 [0.82, 1.06] | 0.282 | 957 | 0.95 [0.78, 1.15] | 0.615 | 1120 | 0.98 [0.82, 1.17] | 0.821 |
| Platelet count | 922 | 0.88 [0.76, 1.03] | 0.134 | 1079 | 0.92 [0.79, 1.06] | 0.271 | 922 | 1.12 [0.93, 1.36] | 0.265 | 1079 | 1.14 [0.95, 1.37] | 0.167 |
| Lymphocytes | 920 | 0.93 [0.81, 1.07] | 0.281 | 1076 | 0.92 [0.81, 1.05] | 0.222 | 920 | 1.04 [0.86, 1.27] | 0.727 | 1076 | 1.04 [0.87, 1.24] | 0.778 |

Notes.
All models in this table are adjusted for age, sex, and comorbidities.

**Supplementary Table 3:** Internally validated discrimination for KCH training sample based on nested repeated cross-validation

| Outcome | Sample | Model | No. features | AUC [95% CI] | Brier score [95% CI] | Sensitivity [95% CI] | Specificity [95% CI] |
| --- | --- | --- | --- | --- | --- | --- | --- |
| 14-day ICU/death<sup>1</sup> | Subset 1: Patients with data on ethnicity (n=960) | NEWS2 + age | 2 | 0.694 [0.667, 0.721] | 0.197 [0.191, 0.204] | 0.848 [0.811, 0.889] | 0.364 [0.324, 0.405] |
|  |  | All features | 9 | 0.728 [0.703, 0.754] | 0.189 [0.183, 0.195] | 0.806 [0.768, 0.845] | 0.496 [0.459, 0.533] |
|  | Subset 2: Excluding nosocomial patients (n=1123) | NEWS2 + age | 2 | 0.721 [0.701, 0.744] | 0.189 [0.184, 0.195] | 0.809 [0.775, 0.842] | 0.474 [0.444, 0.507] |
|  |  | All features | 9 | 0.748 [0.727, 0.770] | 0.183 [0.177, 0.188] | 0.780 [0.743, 0.815] | 0.539 [0.503, 0.571] |
| 3-day ICU/death<sup>2</sup> | All patients (n=1276) | NEWS2 + age | 2 | 0.764 [0.737, 0.794] | 0.114 [0.108, 0.120] | 0.967 [0.871, 1.000] | 0.286 [0.000, 0.461] |
|  |  | All features | 2 | 0.789 [0.763, 0.819] | 0.110 [0.103, 0.116] | 0.856 [0.793, 0.933] | 0.534 [0.475, 0.617] |
|  | Subset 1: Patients with non-missing ethnicity (n=960) | NEWS2 + age | 2 | 0.763 [0.732, 0.796] | 0.114 [0.107, 0.122] | 0.958 [0.857, 1.000] | 0.363 [0.211, 0.473] |
|  |  | All features | 2 | 0.774 [0.740, 0.810] | 0.110 [0.102, 0.118] | 0.838 [0.739, 0.950] | 0.578 [0.429, 0.669] |
|  | Subset 2: Excluding nosocomial patients (n=1123) | NEWS2 + age | 2 | 0.769 [0.738, 0.798] | 0.119 [0.113, 0.126] | 1.000 [0.971, 1.000] | 0.150 [0.000, 0.187] |
|  |  | All features | 2 | 0.796 [0.767, 0.825] | 0.115 [0.108, 0.122] | 0.940 [0.888, 1.000] | 0.343 [0.180, 0.483] |

Notes. <sup>1</sup> Sensitivity/specificity calculated at 30% probability threshold; <sup>2</sup> Calculated at 20% probability threshold.
AUC based on repeated, nested cross-validation (inner loop: 10 folds; outer loop = 10 folds/1000 repeats). Missing values imputed at each outer loop with k-Nearest Neighbours (KNN) imputation.

**Supplementary Table 4:**
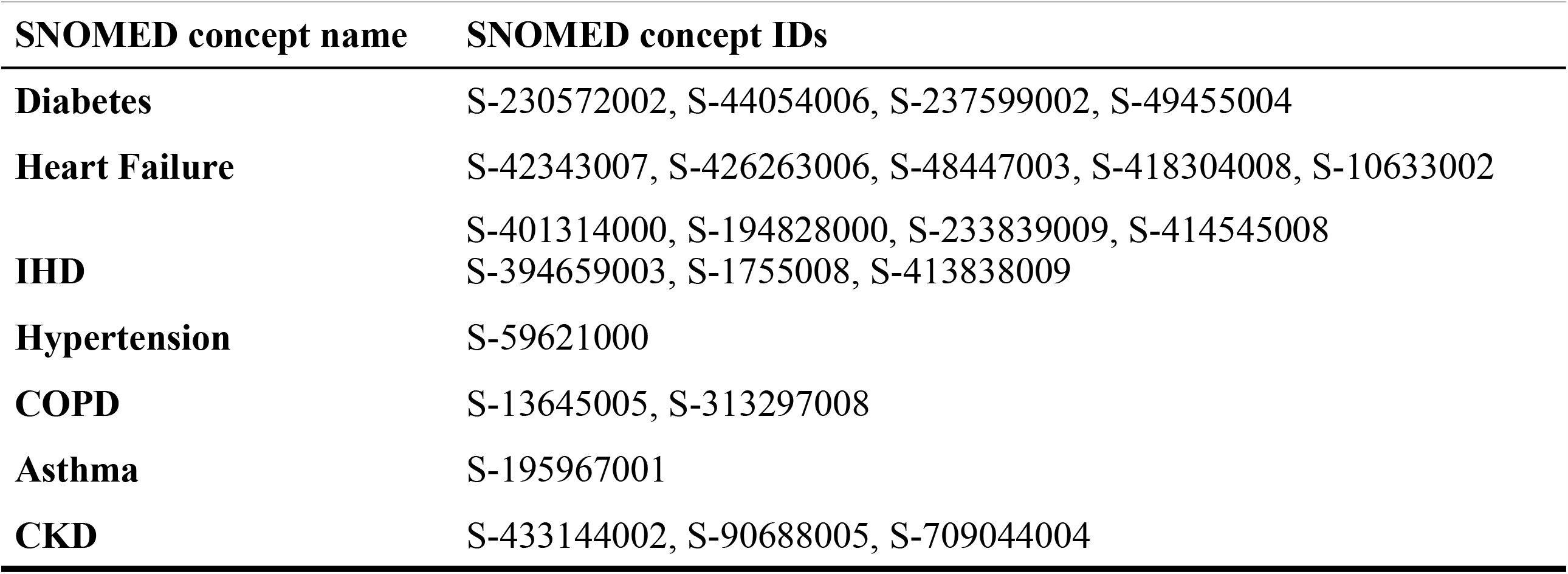
SNOMED terms

**Supplementary Table 5:**
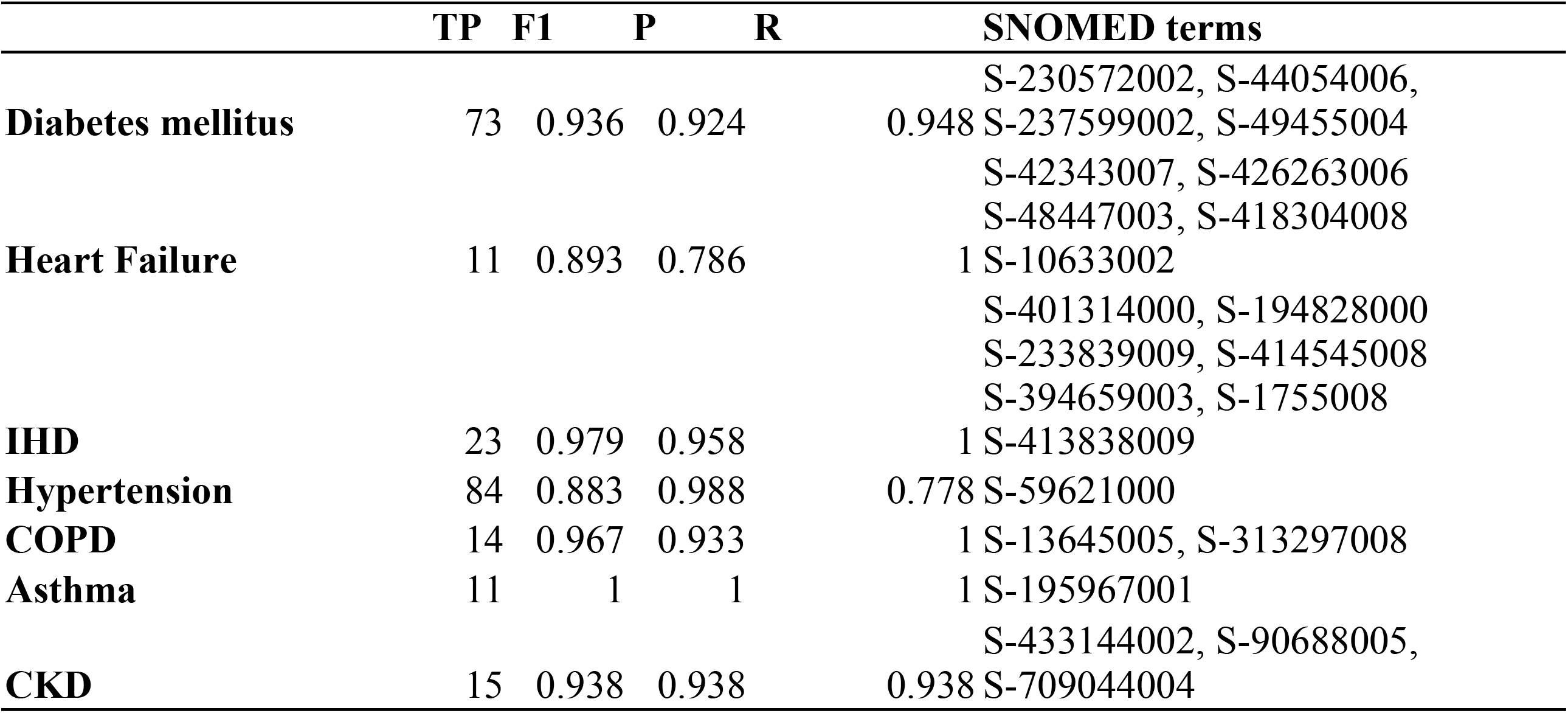
F1, precision and recall for NLP comorbidity detection *MedCATTrainer* was used to collect manual annotations for 146 clinical documents totalling 4343 annotations. Each co-morbidity is defined using one or more SNOMED terms. Predicted true positive labels (TP), precision (P), recall (R), F1-score (F1) are shown for these aggregated concepts. These results only consider entity detection and not meta annotation.

## Footnotes

1 https://github.com/ewancarr/NEWS2-COVID-19

## Notes

### Author Declarations

This project operated under London South East Research Ethics Committee approval (reference 18/LO/2048) granted to the King's Electronic Records Research Interface (KERRI). Given the current context, specific work on COVID19 research was reviewed with a virtually convened 4-expert patient panel input on a virtual committee with Caldicott Guardian oversight. In addition, the KERRI database used in this study was developed according to KCH use of data for research guidelines (https://www.kch.nhs.uk/research/use-of-data-for-research) and patient engagement groups for this dataset have been conducted in 2018-2019.

### Summary of Updates

This revision (i) expands the cohort used for model development to all 1276 patients at KCH; (ii) uses hospital admission (rather than symptom onset) as the index date; (iii) considers shorter-term (3-day) endpoints in sensitivity analyses; (iv) improves reporting of model calibration and clinical utility in validation sites; and (v) increases the number of external sites.

